# Bacterial Contamination and Antimicrobial Susceptibility Patterns of Isolates from Door Handles at a Public University Campus in Uganda: A Cross-Sectional Study

**DOI:** 10.64898/2026.09.02.26362121

**Authors:** Agoe Angella Lilian, Samba David, Ampuriire Annex, Odongo Jackson, Osiime Isaac, Jazira Tumusiime, Okeny Christopher, Benson Musinguzi, Yona Mbalibulha, Simon Peter Rugera

## Abstract

Door handles are among the most frequently touched surfaces in institutional settings and can act as fomites for the transmission of pathogenic and antimicrobial-resistant bacteria, yet contamination on university campuses in low-resource settings remains poorly characterised. We conducted a laboratory-based cross-sectional study to determine the prevalence, bacteriology and antimicrobial susceptibility profile of contaminants on door handles at Mbarara University of Science and Technology (MUST) City Campus, Uganda. Using stratified random sampling across hostels, lecture rooms, toilets, laboratories and offices, 100 door handles were swabbed in the evening using a standardised technique and processed within 24 hours. Isolates were cultured on blood and MacConkey agar, identified by colony morphology, Gram stain and standard biochemical tests, and tested for antimicrobial susceptibility by the Kirby-Bauer disc diffusion method per CLSI guidelines. Of the 100 door handles sampled, 53 (53.0%) yielded bacterial growth, producing 59 isolates across five species, including three cases of polymicrobial contamination. Contamination was highest on toilet handles (68.2%) and lowest on laboratory handles (20.0%). Gram-positive cocci predominated (89.8%), led by *Staphylococcus hyicus* (47.5%) and *Staphylococcus aureus* (40.7%); *Klebsiella pneumoniae* (8.5%), *Hafnia sp*. (1.7%) and *Staphylococcus saprophyticus* (1.7%) were also recovered. Antimicrobial susceptibility testing revealed high resistance to azithromycin (61.0%), tetracycline (44.1%) and penicillin (40.7%), while gentamicin (8.5% resistance) and chloramphenicol (20.3% resistance) retained the greatest activity. Cefixime and ceftriaxone resistance among Gram-negative isolates (tested only on n=6) suggested possible extended-spectrum beta-lactamase production. These findings demonstrate that door handles in a university setting, including non-clinical, community environments, harbour a substantial burden of bacterial contamination and antimicrobial resistance, with implications for infection prevention policy in academic institutions in low- and middle-income countries. Routine surface disinfection, reinforced hand hygiene, and prudent antimicrobial use are recommended, alongside further studies incorporating molecular confirmation and minimum inhibitory concentration testing.

## Introduction

Microbial contamination of high-touch public surfaces is a recognised global health challenge. Shared utilities such as door handles are frequently contaminated with potentially pathogenic bacteria and are increasingly implicated in the indirect transmission of both commensal organisms and antimicrobial-resistant pathogens [1]. University campuses combine high population density with diverse, high-frequency surface contact across residential, academic, sanitary and administrative spaces, yet, despite the frequency of human contact with door handles in this setting, there has been no comprehensive synthesis of contamination rates and microbial profiles specific to university environments in Uganda. Antimicrobial resistance (AMR) is simultaneously recognised as one of the top global public health threats: a 2026 global governance analysis conducted ahead of the WHO Global Action Plan on AMR (2026-2036) update found that, despite measurable improvement in national governance scores, implementation and surveillance continue to lag behind policy design, particularly in the animal and environmental sectors [2,3], with environmental reservoirs, including inanimate surfaces, increasingly implicated in the dissemination of resistant organisms beyond clinical settings. In vitro evidence indicates that many bacterial pathogens can retain culturable viability on inanimate surfaces for periods ranging from hours to several months depending on species, surface material and ambient conditions, supporting the biological plausibility of door handles as vehicles for indirect transmission [4]. We therefore conducted a cross-sectional study to determine the prevalence of bacterial contaminants on door handles and to characterise their antimicrobial susceptibility patterns at Mbarara University of Science and Technology (MUST) City Campus, Uganda, with a view to informing infection prevention practice in academic institutions in a low-resource setting.

## Materials and Methods

### Study design and site

This study employed a laboratory-based cross-sectional design, conducted at MUST City Campus, comprising 120 hostel rooms (male and female), 22 lecture rooms, 19 laboratories (clinical, computer, physics, chemistry and biology), 75 toilets and 70 offices.

### Target population and sampling

The target population comprised all internal and external door handles on high-traffic doors at the campus (103 hostel doors, 24 lecture rooms, 47 toilet doors, 10 laboratories and 30 offices). Sample size was calculated using the Kish-Leslie formula based on a previously reported contamination prevalence of 86% at a comparable Ugandan university [5], with 95% confidence and 5% margin of error, yielding an initial sample size of 185, adjusted for the finite population to a final sample size of 100 door handles. A stratified random sampling technique was used, with the sample proportionally allocated across five strata by room function: hostels (48), toilets (22), offices (14), lecture rooms (11) and laboratories (5); individual door handles were then randomly selected within each stratum. High-traffic external and internal door handles were included; handles on doors adjacent to a sampled high-traffic door were excluded to avoid duplication.

### Sample collection and processing

Sterile swabs pre-moistened with Amie’s transport medium were used, employing a standardised technique over a defined surface area of approximately 20 cm^2^ per handle. Sampling was conducted in the evening (5–6 pm) to capture contamination accumulated after a full day’s use (before end of day cleaning), and swabs were transported to the MUST microbiology laboratory for processing within 24 hours. Each swab was rolled onto blood agar and MacConkey agar and streaked in a zig-zag pattern using a sterile wire loop, then incubated aerobically at 37°C for 18-24 hours. Where growth was non-uniform, single colonies were sub-cultured to obtain pure isolates.

### Bacterial identification and antimicrobial susceptibility testing

Pure isolates were identified by colony morphology, Gram stain reaction, and standard biochemical tests: catalase, coagulase, mannitol salt agar, DNase and novobiocin resistance tests for Gram-positive isolates; and citrate, urease, oxidase, sulphide-indole-motility (SIM), and triple sugar iron (TSI) tests for Gram-negative isolates. Antimicrobial susceptibility testing was performed on all significant isolates by the Kirby-Bauer disc diffusion method on Mueller-Hinton agar, per Clinical and Laboratory Standards Institute (CLSI 2025) guidelines, using an eight-antibiotic panel: penicillin, ceftriaxone, cefixime, azithromycin, ciprofloxacin, gentamicin, tetracycline and chloramphenicol. Ceftriaxone and cefixime were tested on Gram-negative isolates only (n=6).

### Quality control and data analysis

Reference ATCC strains (*Escherichia coli* ATCC 25922, *Staphylococcus aureus* ATCC 25923, *Pseudomonas aeruginosa* ATCC 27853) were used for quality control of biochemical and susceptibility testing. Data were entered into Microsoft Excel 2016, checked for completeness, and exported to SPSS Statistics Version 26 for descriptive analysis (frequencies and percentages).

### Ethical considerations

Ethical approval was obtained from the Mbarara University Faculty Research Committee.(Reference number MUST/MLS/023) Permission to sample was obtained from the Chief Human Resources Officer (Reference number MUST 18/3 and from occupants of rooms whose door handles were swabbed. No personal identifiers were collected on sample or data forms.

## Results

### Prevalence of contamination

Of the 100 door handles sampled across five City campus settings, 53 (53.0%) were contaminated with bacterial pathogens, while 47 (47.0%) showed no growth. Contamination rates varied substantially by location (Table 1), with the highest rates recorded on toilet handles (68.2%, 15/22) and lecture room handles (63.6%, 7/11), followed by hostels (52.1%, 25/48), offices (35.7%, 5/14), and laboratories (20.0%, 1/5).

**Table 1.** Proportion of contaminated door handles by location at MUST City Campus.

| **Location** | **Total sampled (n)** | **Contaminated, %(n/N)** |
| --- | --- | --- |
| Hostels | 48 | 52.1 (25/48) |
| Toilets | 22 | 68.2 (15/22) |
| Laboratories | 5 | 20.0 (1/5) |
| Offices | 14 | 35.7 (5/14) |
| Lecture rooms | 11 | 63.6 (7/11) |
| **Total** | **100** | **53.0 (53/100)** |

### Bacterial species isolated

Culture yielded 59 bacterial isolates from the 53 positive samples, reflecting three cases of polymicrobial contamination; one hostel door handle yielded three concurrent isolates (*Klebsiella pneumoniae, Hafnia sp*., and *Staphylococcus hyicus*). Gram-positive cocci predominated (53/59, 89.8%), with Gram-negative rods accounting for the remainder (6/59, 10.2%). *Staphylococcus hyicus* was the most frequently isolated organism (28/59, 47.5%), followed by *Staphylococcus aureus* (24/59, 40.7%), *Klebsiella pneumoniae* (5/59, 8.5%), and *Hafnia sp*. and *Staphylococcus saprophyticus* (1/59, 1.7% each). *Staphylococcus hyicus* predominated in hostels (14/27, 51.9%) and offices (3/6, 50.0%); *Staphylococcus aureus* was most common in lecture rooms (4/7, 57.1%) and toilets (7/18, 38.9%); *Klebsiella pneumoniae* was isolated mainly from toilets (3/5, 60.0%).

### Antimicrobial susceptibility patterns

Among the 59 isolates tested, azithromycin showed the highest resistance (36/59, 61.0%), followed by tetracycline (26/59, 44.1%) and penicillin (24/59, 40.7%). Ciprofloxacin resistance was observed in 25.4% (15/59) of isolates and chloramphenicol resistance in 20.3% (12/59). Gentamicin was the most effective agent, with resistance in only 8.5% (5/59) of isolates. Among the six Gram-negative isolates tested against ceftriaxone and cefixime, 50.0% (3/6) were resistant to cefixime and 16.7% (1/6) to ceftriaxone. 6 (10.1%) out of the 59 isolates were not tested using penicillin since they were gram negative isolates (Table 2).

**Table 2.** Antimicrobial susceptibility of bacterial isolates (N=59). *Ceftriaxone and cefixime were tested only on Gram-negative isolates (n=6); remaining isolates recorded as not done.

| Antibiotic | Sensitive, %(n) | Intermediate, %(n) | Resistant, %(n) |
| --- | --- | --- | --- |
| Penicillin | 47.5 (28) | 1.7 (1) | 40.7 (24) |
| Azithromycin | 39.0 (23) | 0 | 61.0 (36) |
| Tetracycline | 55.9 (33) | 0 | 44.1 (26) |
| Ciprofloxacin | 39.0 (23) | 35.6 (21) | 25.4 (15) |
| Chloramphenicol | 69.5 (41) | 10.2 (6) | 20.3 (12) |
| Gentamicin | 71.2 (42) | 20.3 (12) | 8.5 (5) |
| Ceftriaxone* | 6.8 (4) | 1.7 (1) | 1.7 (1) |
| Cefixime* | 3.4 (2) | 1.7 (1) | 5.1 (3) |

## Discussion

This study found that 53.0% of door handles sampled across a Ugandan university campus were contaminated with bacteria, a finding consistent with the broader pattern of substantial fomite contamination reported across sub-Saharan African institutional settings and other low- and middle-income countries [1,6], including a 2025 comparative survey of hospital and community door handles in Nigeria that recorded contamination rates as high as 100% on university toilet handles and identified them as the principal reservoir of multidrug-resistant isolates [7]. The recovery of 59 isolates from 53 positive samples, including three instances of polymicrobial contamination, underscores that a single contact event on a communal door handle can transfer multiple organisms with differing virulence and resistance profiles, reinforcing the value of considering such surfaces as composite rather than single-organism infection risks.

Contamination was highest on toilet (68.2%) and lecture room (63.6%) handles, a pattern consistent with reports from Nigeria linking elevated contamination to high-density, low-intervening-hygiene traffic in these settings [7,8]. The elevated toilet contamination likely reflects fecal-oral transfer from imperfect post-use hand hygiene and aerosolization during flushing [8], while the high contamination of lecture room handles likely reflects sequential high-frequency touching without intervening hand hygiene throughout the academic day. This is consistent with recent evidence from a Nigerian university campus in which hand hygiene knowledge among undergraduate students was found to be poor despite generally positive attitudes, with forgetfulness and inadequate handwashing facilities cited as key barriers to practice [9]. Lower contamination in offices and laboratories is consistent with less frequent use and, in laboratories, adherence to routine hygiene practices such as glove use and surface decontamination [10].

Gram-positive cocci, principally *Staphylococcus species*, accounted for 89.8% of isolates, consistent with their known origin in human skin, nasal and respiratory secretions and their established predominance on environmental surfaces in educational and healthcare settings [6,11]. *Staphylococcus hyicus*, the most prevalent isolate (47.5%), is classically an animal pathogen but is increasingly reported as an environmental coloniser capable of transient human carriage [12,13], and its widespread recovery here suggests a stable environmental reservoir on campus surfaces rather than a transient event. *Staphylococcus aureus* (40.7%) was the most clinically significant organism identified, given its established role in skin and soft tissue infection, bacteraemia and toxin-mediated disease [14]; the substantial resistance observed in *S. aureus* isolates to azithromycin (66.7%), tetracycline (50.0%) and chloramphenicol (33.3%) raises concern for circulating multidrug-resistant strains, although methicillin resistance was not directly tested. A 2025 survey of high-touch hospital surfaces, including door handles, in Cameroon similarly found *S. aureus* isolates to be highly susceptible to gentamicin despite resistance to other agents, a pattern consistent with our own findings [15].

*Klebsiella pneumoniae*, a WHO priority pathogen for antimicrobial resistance [16], was recovered mainly from toilet handles; resistance to cefixime (40.0%) and ceftriaxone (20.0%) among these isolates, together with predominantly intermediate gentamicin susceptibility, is suggestive of possible extended-spectrum beta-lactamase (ESBL) production, consistent with reports from Uganda and Nigeria [17,18]. The single *Hafnia sp*. isolate, though rare, is notable given its recognised role as an emerging opportunistic pathogen [19], and its co-isolation with *K. pneumoniae* and *S. hyicus* from a single hostel handle exemplifies the polymicrobial contamination risk documented in this study.

Across the antibiotic panel, resistance was highest to azithromycin, tetracycline and penicillin, while gentamicin and chloramphenicol retained the greatest activity, mirroring global trends of widespread macrolide and beta-lactam resistance alongside relatively preserved aminoglycoside activity in comparable environmental surveillance [20,21]. These findings support gentamicin as a priority empirical agent for infections potentially arising from environmental exposure of this kind, while highlighting azithromycin, tetracycline and penicillin as poor empirical choices in this setting.

## Conclusion

This study demonstrates substantial bacterial contamination of door handles at a Ugandan university campus, with over half of sampled surfaces yielding bacterial growth dominated by *Staphylococcus hyicus* and *Staphylococcus aureus*, alongside the clinically significant, potentially ESBL-producing pathogen *Klebsiella pneumoniae*. High resistance to commonly used antibiotics, particularly azithromycin, penicillin and tetracycline, was observed, while gentamicin and chloramphenicol remained comparatively effective. These results support the implementation of regular disinfection for frequently touched surfaces, enhanced handwashing protocols, and cautious antibiotic use within university settings. Furthermore, they emphasize the importance of subsequent research utilising molecular validation for species identification and methicillin resistance, alongside minimum inhibitory concentration analysis, to more accurately profile the resistance impact of environmental strains in academic institutions within low-resource settings.

## Limitations

Bacterial identification relied on conventional biochemical methods without molecular confirmation, which may not reliably differentiate closely related species. Antimicrobial susceptibility testing used the disc diffusion method, which does not provide minimum inhibitory concentration values for more precise resistance characterisation. Analysis was restricted to descriptive statistics (frequencies and percentages); formal inferential comparison of contamination and resistance proportions across locations was not performed, and should be considered in future studies with adequately powered strata.

This study considered only mesophilic bacterial organisms (defined as those growing best at moderate temperatures, typically between 20°C and 45°C), and did not include potentially pathogenic anaerobic organisms such as *Clostridioides difficile* and *Clostridium perfringens*.

## Acknowledgments

The authors thank the administration of Mbarara University of Science and Technology for permission to conduct this study, and the Department of Medical Laboratory Science for providing the academic and laboratory resources necessary for its completion.

## Author Contributions

Conceptualization: Samba David, Ampuriire Annex, Odongo Jackson, Osiime Isaac, Agoe Angella Lilian, Yona Mbalibulha, Simon Peter Rugera. Data curation, Formal analysis, Investigation: Samba David, Ampuriire Annex, Odongo Jackson, Osiime Isaac, Agoe Angella Lilian. Supervision: Yona Mbalibulha, Simon Peter Rugera. Writing – original draft: Samba David, Ampuriire Annex, Odongo Jackson, Osiime Isaac, Agoe Angella Lilian. Draft review and polishing: Jazira Tumusiime, Okeny Christopher, Musinguzi Benson. Writing – final review & editing: Benson Musinguzi, Yona Mbalibulha, Simon Peter Rugera.

## Data Availability

The datasets generated and analysed during the current study are available from the corresponding author on reasonable request.

